# Effects of the COVID-19 Pandemic on Individuals with Fibromyalgia – a Systematic Scoping Review Protocol

**DOI:** 10.1101/2023.01.27.23284843

**Authors:** Tali Sahar, Ali Jalali, Sylvie Toupin, Maria Verner, Sabrina Mitrovic, Amir Minerbi, Yoram Shir, Mary-Ann Fitzcharles, M. Gabrielle Pagé

## Abstract

**Objective:** The objectives of this review are to systematically search databases and identify studies that examined the effects of COVID-19 pandemic on symptomatology of adults who had fibromyalgia prior to the pandemic, in order to map the existing knowledge and identify knowledge gaps.

**Introduction:** The COVID-19 pandemic has affected people worldwide in multiple ways. Some suffered infection of varying severity and many experienced stressors associated with quarantine restrictions, lockdowns, and the consequences of social distancing. An initial literature search indicates that the pandemic had different and sometime contradicting effects on individuals with fibromyalgia; while some people experienced worsening of symptoms, others reported symptom relief because of the reduced pace and demands of daily life.

**Inclusion criteria:** Any studies that explored the experience of adults with fibromyalgia syndrome during the COVID-19 pandemic. We will review only studies with participants who were diagnosed with fibromyalgia prior to the pandemic.

**Methods:** Following a pilot search, we developed a full search strategy for Medline, Embase, CINAHL and PsycInfo. The reference list of all included sources of evidence will be screened for additional studies. Sources of unpublished studies to be searched: clinical trial.gov, OPENGREY.EU and MedRxiv. Studies in any language will be included. Abstracts will be screened for inclusion by two reviewers. Similarly, two independent reviewers will systematically extract the data from the included articles. Disagreements in any stage will be resolved through consensus. The results will be presented in tables and will be accompanied by a narrative analysis.

## Introduction

### Rationale

The COVID-19 pandemic has affected people worldwide in multiple ways. Some were infected with the virus with varying severity of illness, others were affected by illness within families, and many experienced stressors associated with quarantine restrictions, lockdowns, and the consequences of social distancing^1^. Patients with chronic illnesses experienced stressors like the general population, at large, but with the added burden of living with a chronic illness and changes that may have occurred in healthcare delivery. Experts within the healthcare community raised concerns that the pandemic would adversely affect patients with chronic noncancer pain (CNCP)^2,3^. Indeed, a large Pan-Canadian survey has supported these concerns^4^. There were, however, reports early in the pandemic that patients with CNCP did not experience symptom exacerbations. The pandemic may have been advantageous for some due to the reduced pace of daily life that was imposed by social distancing and the resulting restrictions^5,6^.

Fibromyalgia syndrome (FMS) is a prevalent pain condition that is known to be sensitive to various stressors, with patients reporting exacerbation of symptoms related to psychological and environmental stressors. In this context, it has been questioned whether the symptoms of FMS were adversely affected by the pandemic by such parameters as personal infection, stressors related to family infection and changes in healthcare access or working environment, or conversely whether some experienced symptom relief. An initial literature search provided contradicting evidence supporting both hypotheses, with some studies reporting worsening of pain and FM symptoms,^7^ whereas others reported no change or improvement of symptoms^8^. In a study of 32 Italian patients with FMS who were contacted at the end of the lockdown period, Cavalli et al.^9^ reported that while the median Revised Fibromyalgia Impact Questionnaire (FIQR) scores of patients did not change before and after the lockdown, two thirds had worsening of symptoms with higher scores, while one third had either no change or improvement. Understanding the diverse effects of the pandemic on individuals with FMS will better inform healthcare professionals in management of these patients when exposed to stressors. Strategies and measures that were perceived as helpful and could have led to improvement may be further examined with the opportunity to be integrated into routine clinical care. Furthermore, we hope that our review will help direct researchers towards patient focused studies that are clinically relevant^10^.

We will systematically review studies that explored the effects of the COVID-19 pandemic on individuals with FMS. Our aim is to explore and synthesize current knowledge, with the objective to obtain clinically significant insights, as well as to identify research gaps. These aims are in line with the definition of a “scoping review” which is to ‘map the literature on a particular topic or research area and provide an opportunity to identify key concepts; gaps in the research; and types and sources of evidence to inform practice, policymaking, and research^11^. We also used the online decision tool “right review” (*Previously known as “What Review is Right for You?”* To assess which type of review to choose https://whatreviewisrightforyou.knowledgetranslation.net/‘

A preliminary search of MEDLINE, the Cochrane Database of Systematic Reviews and Joanna Briggs Institute (*JBI) Evidence Synthesis* was conducted and no current or underway systematic reviews or scoping reviews on the topic were identified.

### Objectives

The objectives of this review are to systematically search databases and identify studies that examined the effects of the Covid-19 pandemic on symptomatology of adults who had fibromyalgia prior to the pandemic, and to systematically extract and evaluate those studies. We aim to map the existing knowledge and identify knowledge gaps.

## Review question

What is known about the experiences of individuals with FMS during the COVID-19 pandemic. Specifically: How did the pandemic influence adults with FMS regarding:

- Pain
- Quality of life
- Sleep
- Mental health
- Work status
- FMS symptoms other than pain
- Changes in pharmacological, psychological, and physical treatments and accessibility to relevant therapies.

## Eligibility criteria

### Participants, Concept and Context

Any studies that explored the experience of adults with FMS during the COVID-19 pandemic and the impact of several aspects of the pandemic, such as: lockdowns, changes in healthcare accessibility, remote work, quarantines. We will exclude any studies of participants who did not have a diagnosis of FMS prior to the pandemic. Studies of long-covid and fibromyalgia-like symptomatology post Covid-19 infection, will be excluded. In mixed-population studies (e.g. studies of individuals over 16 years, or studies that included individuals with pain diagnoses other than FMS) will be included if data are presented separately for each population. When data for mixed population is not presented separately – we will consider inclusion if mor than 75% of the participants will be considered for inclusion if more than 75% of the participants had FMS.

### Types of Sources

This scoping review will consider any study presenting original data. Any study design, including analytical observational studies including prospective and retrospective cohort studies, case-control studies and analytical cross-sectional studies will be considered for inclusion. This review will also consider descriptive observational study designs including case series, individual case reports and descriptive cross-sectional studies for inclusion. Qualitative studies will also be considered. In addition, systematic reviews that meet the inclusion criteria will also be considered. Opinion papers will not be considered for inclusion in this scoping review.

## Methods

The proposed scoping review will be conducted in accordance with the PRISMA Extension for Scoping Reviews PRISMA-ScR^12^ and the JBI methodology for scoping reviews^13^

### Search strategy

The search strategy will aim to locate both published and unpublished studies. An initial limited search of MEDLINE was undertaken to identify articles on the topic. The text words contained in the titles and abstracts of relevant articles, and the index terms used to describe the articles were used to develop a full search strategy for Medline, Embase, CINAHL and PsycInfo (see Appendix #). The reference list of all included sources of evidence will be screened for additional studies. Sources of unpublished studies/grey literature to be searched: clinicaltrials.gov, OPENGREY.EU and MedRxiv.

Members of our team can interpret studies in the following languages: English, French, Spanish, Italian, Hebrew, Arabic, Farsi, German, Portuguese and Russian. We will strive to obtain official translation of studies from other languages, if needed.

### Study/Source of Evidence selection

Following the search, all identified citations will be collated and uploaded into Covidence (Clarivate Analytics, PA, USA, (2020 edition)) and duplicates removed. Following a pilot test, titles and abstracts will be screened by two independent reviewers (AJ and TS) for compatibility with the review inclusion criteria. Potentially relevant texts will be retrieved in full. The full text of selected citations will be assessed in detail against the inclusion criteria by two independent reviewers (ST and SM). Reasons for the exclusion of sources not meeting the inclusion criteria will be recorded and reported in the scoping review. Any disagreements that arise between the reviewers at each stage of the selection process will be resolved through discussion, or by consulting an additional reviewer/s. The results of the search and the study inclusion process will be reported in full in the final scoping review and presented in a Preferred Reporting Items for Systematic Reviews and Meta-analyses extension for scoping review (PRISMA-ScR) flow diagram^12^

### Data Extraction

Data will be extracted from sources included in the scoping review by two or more independent reviewers using a dedicated data extraction template developed by the reviewers (MV, AJ, GP, TS, ST). The data extracted will include specific details about the participants, concept, context, study methods and key findings relevant to the review questions.

A draft extraction form is provided (see Appendix XX). The preliminary data extraction template will be modified and revised as necessary during the process of data extraction from each included evidence source. Modifications will be detailed in the scoping review. Any disagreements that arise between the reviewers will be resolved through discussion, or with an additional reviewer/s. If required, authors of relevant papers will be contacted to request missing or additional data.

### Data Analysis and Presentation

The data will be presented in a table with relevant diagrams and/or graphs, in case these provide a clearer view. A narrative summary will accompany the tabulated presentation, describing how the results relate to the review objective and questions.

## Supporting information

Search Strategy

Data extraction Form

## Data Availability

All data produced in the present study are available upon reasonable request to the authors

## Acknowledgements

The authors gratefully acknowledge Alex Amar (MLIS), from the McGill University Health Centre (MUHC) Medical Libraries, for the kind and abled assistance with the search protocol and methodology.

