## Supplementary material for "Effects of the COVID-19 Pandemic on Individuals with Fibromyalgia – a Systematic Scoping Review Protocol": Search Strategy

**MUHC Libraries / Bibliotheques CUSM**

| | |
|---|---|
| Literature search | MGH-1481 |
| Conducted by | Alex Amar, MLIS |
| At | McGill University Health Centre Medical Libraries |
| For | Tali Sahar, MD |
| On the topic of | COVID-19 in patients with Fibromyalgia |
| On | 7/5/2022 |
| Delivered on | 7/6/2022 |
| Notes: | COVID search terms modified from Canadian Agency for Drugs and Technologies in Health - https://covid.cadth.ca/literature-searching-tools/cadth-covid-19-search-strings/ |
| Concerning full text access: | Your librarian can assist you in retrieving the full text of articles.<br><br>The MUHC Libraries cover the fees for most interlibrary loans. However, charges may apply for high volume requests or difficult-to-obtain items. You will be informed of any such charges before your request is processed. |
| Please consider providing feedback on this literature search: https://www.surveymonkey.com/r/lit-search-N1 | |

### Search Strategies

#### Medline

| Ovid MEDLINE(R) and Epub Ahead of Print, In-Process, In-Data-Review & Other Non-Indexed Citations and Daily <1946 to July 01, 2022> | | |
|---|---|---|
| 1 | COVID-19/ | 171537 |
| 2 | exp COVID-19 Testing/ | 9060 |
| 3 | exp COVID-19 Vaccines/ | 13686 |
| 4 | exp SARS-CoV-2/ | 130377 |
| 5 | (coronavirus/ or betacoronavirus/ or coronavirus infections/) and (disease outbreaks/ or epidemics/ or pandemics/) | 40138 |
| 6 | (nCoV* or 2019nCoV or 19nCoV or COVID19* or COVID or SARS-COV-2 or SARSCOV-2 or SARS-COV2 or SARSCOV2 or SARS coronavirus 2 or Severe Acute Respiratory Syndrome Coronavirus 2 or Severe Acute Respiratory Syndrome Corona Virus 2).ti,ab,kf,nm,ot,ox,rx,px. | 261729 |
| 7 | ((new or novel or "19" or "2019" or "2020" or Wuhan or Hubei or China or Chinese) adj3 (coronavirus* or corona virus* or betacoronavirus* or CoV or HCoV)).ti,ab,kf,ot. | 72832 |
| 8 | (longCOVID* or postCOVID* or postcoronavirus* or postSARS*).ti,ab,kf,ot. | 43 |
| 9 | ((coronavirus* or corona virus* or betacoronavirus*) adj3 (pandemic* or epidemic* or outbreak* or crisis)).ti,ab,kf,ot. | 12832 |
| 10 | ((Wuhan or Hubei) adj5 pneumonia).ti,ab,kf,ot. | 398 |
| 11 | or/1-10 | 273331 |
| 12 | fibromyalg*.tw,kf. | 11840 |
| 13 | Fibromyalgia/ | 9462 |
| 14 | (fibro or fibromyalg*).af. | 19410 |
| 15 | or/12-14 | 19410 |
| 16 | 11 and 15 | 74 |

#### Embase

| Embase <1974 to 2022 July 01> | | |
|---|---|---|
| 1 | sars-related coronavirus/ | 512 |
| 2 | (coronavirinae/ or betacoronavirus/ or coronavirus infection/) and (epidemic/ or pandemic/) | 10806 |
| 3 | (nCoV* or 2019nCoV or 19nCoV or COVID19* or COVID or SARS-COV-2 or SARSCOV-2 or SARS-COV2 or SARSCOV2 or SARS coronavirus 2 or Severe Acute Respiratory Syndrome Coronavirus 2 or Severe Acute Respiratory Syndrome Corona Virus 2).ti,ab,kw,hw,ot. | 282432 |
| 4 | ((new or novel or "19" or "2019" or Wuhan or Hubei or China or Chinese) adj3 (coronavirus* or corona virus* or betacoronavirus* or CoV or HCoV)).ti,ab,kw,hw,ot. | 245293 |
| 5 | (longCOVID* or postCOVID* or postcoronavirus* or postSARS*).ti,ab,kw,hw,ot. | 118 |
| 6 | ((coronavirus* or corona virus* or betacoronavirus*) adj3 (pandemic* or epidemic* or outbreak* or crisis)).ti,ab,kw,ot. | 14930 |
| 7 | ((Wuhan or Hubei) adj5 pneumonia).ti,ab,kw,ot. | 459 |
| 8 | or/1-7 | 305232 |
| 9 | limit 8 to yr="2019 -Current" | 303891 |
| 10 | (fibro or fibromyalg*).ti,ab,kf,hw,ot,dq. | 33145 |
| 11 | fibromyalgia/ | 23272 |
| 12 | or/10-11 | 33145 |
| 13 | 9 and 12 | 220 |

#### CINAHL

| Tuesday, July 05, 2022 3:30:39 PM | | |
|---|---|---|
| S1 | (MH "COVID-19+") OR (MH "Post-Acute COVID-19 Syndrome") OR (MH "COVID-19 Testing") OR (MH "COVID-19 Vaccines") OR (MH "COVID-19 Pandemic") OR (MH "SARS-CoV-2") | 61,378 |
| S2 | TI ( nCoV* or 2019nCoV or 19nCoV or COVID19* or COVID or SARS-COV-2 or SARSCOV-2 or SARS-COV2 or SARSCOV2 or SARS coronavirus 2 or Severe Acute Respiratory Syndrome Coronavirus 2 or Severe Acute Respiratory Syndrome Corona Virus 2 ) OR AB ( nCoV* or 2019nCoV or 19nCoV or COVID19* or COVID or SARS-COV-2 or SARSCOV-2 or SARS-COV2 or SARSCOV2 or SARS coronavirus 2 or Severe Acute Respiratory Syndrome Coronavirus 2 or Severe Acute Respiratory Syndrome Corona Virus 2 ) | 88,211 |
| S3 | TI ( ((new or novel or "19" or "2019" or Wuhan or Hubei or China or Chinese) N3 (coronavirus* or corona virus* or betacoronavirus* or CoV or HCoV)) ) OR AB ( ((new or novel or "19" or "2019" or Wuhan or Hubei or China or Chinese) N3 (coronavirus* or corona virus* or betacoronavirus* or CoV or HCoV)) ) | 15,671 |
| S4 | TI ( longCOVID* or postCOVID* or postcoronavirus* or postSARS* ) OR AB ( longCOVID* or postCOVID* or postcoronavirus* or postSARS* ) | 11 |
| S5 | TI ( ((coronavirus* or corona virus* or betacoronavirus*) N3 (pandemic* or epidemic* or outbreak* or crisis)) ) OR AB ( ((coronavirus* or corona virus* or betacoronavirus*) N3 (pandemic* or epidemic* or outbreak* or crisis)) ) | 8,456 |
| S6 | TI ( ((Wuhan or Hubei) N5 pneumonia) ) OR AB ( ((Wuhan or Hubei) N5 pneumonia) ) | 145 |
| S7 | S1 OR S2 OR S3 OR S4 OR S5 OR S6 | 103,929 |
| S8 | (MH "Fibromyalgia") | 6,243 |
| S9 | TI ( fibro or fibromyalg* ) OR AB ( fibro or fibromyalg* ) | 7,113 |
| S10 | S8 OR S9 | 8,321 |
| S11 | S7 AND S10 | 22 |

#### PsycInfo

| APA PsycInfo <1987 to June Week 4 2022> | | |
|---|---|---|
| 1 | exp COVID-19/ | 10514 |
| 2 | exp Coronavirus/ | 12711 |
| 3 | exp Pandemics/ | 8881 |
| 4 | severe acute respiratory syndrome/ | 316 |
| 5 | (nCoV* or 2019nCoV or 19nCoV or COVID19* or COVID or SARS-COV-2 or SARSCOV-2 or SARS-COV2 or SARSCOV2 or SARS coronavirus 2 or Severe Acute Respiratory Syndrome Coronavirus 2 or Severe Acute Respiratory Syndrome Corona Virus 2).af. | 24761 |
| 6 | ((new or novel or "19" or "2019" or "2020" or Wuhan or Hubei or China or Chinese) adj3 (coronavirus* or corona virus* or betacoronavirus* or CoV or HCoV)).tw,id. | 3378 |
| 7 | (longCOVID* or postCOVID* or postcoronavirus* or postSARS*).tw,id. | 2 |
| 8 | ((coronavirus* or corona virus* or betacoronavirus*) adj3 (pandemic* or epidemic* or outbreak* or crisis)).tw,id. | 1344 |
| 9 | 1 or 2 or 3 or 4 or 5 or 6 or 7 or 8 | 25855 |
| 10 | fibromyalgia/ | 2233 |
| 11 | (fibro or fibromyalg*).tw,id. | 3639 |
| 12 | 10 or 11 | 3663 |
| 13 | 9 and 12 | 7 |

#### Guide to search terms

Legends for Medline (Ovid), Embase (Ovid) & CINAHL (Ebsco) are available on our website:
http://www.muhclibraries.ca/Documents/Database_Legends.pdf
