## Supplementary material for "Effects of the COVID-19 Pandemic on Individuals with Fibromyalgia – a Systematic Scoping Review Protocol": Data extraction Form

PREVIEW

### General information

**Study ID**

**Title**

Title of paper / abstract / report that data are extracted from

**Lead author contact details**

**Country in which the study conducted**

1. United States
2. UK
3. Canada
4. Australia
5. Other

**Notes**

### Characteristics of included studies

#### Methods

**Aim of study**

**Study design**

1. Randomised controlled trial
2. Non-randomised experimental study
3. Cohort study
4. Cross sectional study
5. Case control study
6. Systematic review
7. Qualitative research
8. Prevalence study
9. Case series
10. Case report
11. Diagnostic test accuracy study
12. Clinical prediction rule
13. Economic evaluation
14. Text and opinion
15. Other

**Date of publication**

**Dates of study**

1. Acute (3-5/2020)
2. Sub-acute (5-10/2020)
3. Prolonged phase of the pandemic

**Study funding sources**

**Possible conflicts of interest for study authors**

**Possible conflicts of interest for study authors**

1. No
2. Yes

3. Unclear

#### Participants

**Population description**

**Inclusion criteria**

**Exclusion criteria**

**Method of recruitment of participants**

1. Phone
2. Mail
3. Clinic patients
4. Voluntary
5. Other

**Total number of participants**

**Pre-pandemic cohort**

1. Yes
2. No

**If yes: Same cohort reassessed?**

1. Yes
2. No

**Control group**

1. ○ Yes
2. ○ No

**Number of control participants**

**Percent of females**

#### Fibromyalgia

**How Fibromyalgia diagnosis was done**

**Questionnaires**

1. ☐ SSS
2. ☐ FIQ
3. ☐ WPI
4. ☐ Other

**Other: name other questionnaires used**

**Baseline Population Characteristics**

| | Intervention 1 | Intervention 2 | Overall |
|---|---|---|---|
| **Characteristic 1** | | | |
| **Characteristic 2** | | | |
| **Characteristic 3** | | | |
| **Characteristic 4** | | | |

**Intervention and Comparisons**

| | Intervention 1 | Intervention 2 |
|---|---|---|
| **Characteristic 1** | | |
| **Characteristic 2** | | |
| **Characteristic 3** | | |
| **Characteristic 4** | | |

**Outcome table (Acute period of pandemic)**

| | Mean | SD |
|---|---|---|
| **Intervention 1** | | |
| **Intervention 2** | | |

**Outcome table (Prolonged period of pandemic)**

| | Mean | SD |
|---|---|---|
| **Intervention 1** | | |
| **Intervention 2** | | |
